# Using Machine-Learning Techniques to Identify Responders vs. Non-responders in Randomized Clinical Trials

**DOI:** 10.1101/2020.11.21.20232041

**Authors:** Vasiliki Nikolodimou, Paul Agapow

## Abstract

Despite the expectation of heterogeneity in therapy outcomes, especially for complex diseases like cancer, analyzing differential response to experimental therapies in a randomized clinical trial (RCT) setting is typically done by dividing patients into responders and non-responders, usually based on a single endpoint. Given the existence of biological and patho-physiological differences among metastatic colorectal cancer (mCRC) patients, we hypothesized that a data-driven analysis of an RCT population outcomes can identify sub-types of patients founded on differential response to Panitumumab - a fully human monoclonal antibody directed against the epidermal growth factor receptor.

Outcome and response data of the RCT population were mined with heuristic, distance-based and model-based unsupervised clustering algorithms. The population sub-groups obtained by the best performing clustering approach were then examined in terms of molecular and clinical characteristics. The utility of this characterization was compared against that of the sub-groups obtained by the conventional responders analysis and then contrasted with aetiological evidence around mCRC heterogeneity and biological functioning.

The Partition around Medoids clustering method results into the identification of seven sub-types of patients, statistically distinct from each other in survival outcomes, prognostic biomarkers and genetic characteristics. Conventional responders analysis was proven inferior in uncovering relationships between physical, clinical history, genetic attributes and differential treatment resistance mechanisms.

Combined with improved characterization of the molecular subtypes of CRC, applying Machine Learning techniques, like unsupervised clustering, onto the wealth of data already collected by previous RCTs can support the design of further targeted, more efficient RCTs and better identification of patient groups who will respond to a given intervention.

## 1 Introduction

Colorectal cancer (CRC) is responsible for over half a million of deaths globally every year, ranking as the 3rd leading cause of cancer death [1]. Although current therapy practice results in prolongation of median survival time for metastatic patients, this applies to only a third of patients as 65-75% of patients die of the disease due to innate resistance to treatment [2]. Evidence from Randomized Control Trials (RCTs) supports the idea that advanced CRC patients with mutated RAS/BRAF gene status are “non-responders”, meaning they do not benefit from treatments based on epidermal growth factor receptor (EGFR) directed monoclonal antibodies (mAbs), like Panitumumab [3,4,5,6]. Although testing for RAS status in patients who qualify for EGFR inhibitors is nowadays almost standard practice [7,8], considering the response rates to current treatments, it is imperative to scrutinize the mechanisms of resistance and understand predictive biomarkers to develop more efficient drugs for CRC. Successful treatment tailored to specific patients requires as a prerequisite accurate patient sub-grouping, that is identification of distinct patient types based on their differing response to treatment agents. Although a common foundation of CRC sub-typing exists today [9], it remains to be refined with further omics data and drug intervention clinical outcomes to further elucidate our understanding of tumour biology and its link to response heterogeneity.

Understanding heterogeneity of outcomes is bounded by the technical approach by which *differential outcomes and response* are being defined and detected. In the RCT setting, the outcome of an experimental treatment (vs. standard of care or placebo) is typically defined based on the mean difference of the outcome variable(s) - tumour size as an example - between the two arms (experimental vs. control). An alternative way of defining the outcome is comparing the percentage of patients who “respond” to the treatment in the two arms - percentage of patients in the experimental drug arm who achieved a target reduction of tumour size vs. that in the control arm - as an example. This latter technique is known as “responders analysis” [10]. Further analysis suggests that despite its advantages, limitations also apply to the responders analysis approach. An exhaustive review of the limitations is out of scope of this study, however, some key areas of critique are:

- The significance of the physiological response for an individual needs to be studied while considering variations of response within and across individuals [11].
- Dichotomization of a continuous variable comes at the cost of information loss which results into loss of statistical power [12].
- Responders analysis is in principle based on an arbitrary cut-off set by clinical experts in each field. Even when the division is based on a consistent and clinically correct threshold, it can still fail in discerning between patients who achieve an exceptional benefit to the response vs. ones who just make it past the cut-off [13].
- Lastly, the true treatment effect cannot be studied without counterfactuals: how a patient randomized to the experimental drug would have done if they had also been left untreated or and how patients randomized to the control arm, would have reacted, should they be treated with the experimental drug. In that sense, responders analysis is just a way to bypass the true limitation of a parallel trial design rather than a way to treat the underlying problem [14].

Many alternative approaches have been proposed to circumvent the pitfalls of responders analysis [15], [16], [17,18]. Model-based responder prediction analysis has also been proposed as a solution to the limitations of traditional responders analysis. Some of the most recent and promising response modeling techniques propose linear mixed models with interaction terms of treatment with predicting variables [19], probabilistic models for the response outcome [20], construction of treatment-specific algorithm to identify patient profiles associated to beneficial treatment effects [21,22], cross-validation techniques to predict treatment causal effects based on experimental studies results [23] and prediction of individualized treatment effects based on Bayesian non-parametric modelling [24]. Additionally, the rise of Machine Learning (ML) and big data sciences has acted as a catalyst in scientists’ ability to select effective genes and biomarkers that accurately predict or explain chemotherapy responses [25,26].

Although the use of ML techniques is therefore not new to the analysis of treatment subgroups, limited studies have leveraged such approaches to characterize response *per se* and mine the “truth in the data” so to identify patients belonging into distinct treatment effect subgroups. Our hypothesis is that such an algorithmic approach can overcome the limitations of traditional responders analysis and provide a framework for patient characterization based on differential response to a treatment. More specifically, engineering the raw response data collected in a RCT and mining those via unsupervised clustering algorithms can identify patterns of treatment response that shall correspond to distinct patient subgroups. The profile of those response-distinct subgroups can then pave the way for the clinical and biological characterization of those differential response patterns. To demonstrate that, this study examines the application of different ML algorithms on permutations of data collected for the response of metastatic CRC (mCRC) patients to a treatment as part of a RCT and compares the utility of their results to those of the traditional responders analysis in support of aetiological hypotheses around treatment effectiveness scenarios.

## 2 Methodology

### 2.1 Analysis Framework

This study is based on data collected for a phase III RCT - which investigated Panitumumab, an antiEGFR (epidermal growth factor receptor) drug, plus Best Supportive Care (BSC) compared with BSC alone in patients with mCRC. The results of the study have been previously published [27,28]. The current study is focused in re-examining the collected data to investigate differential response to treatment via the use of ML unsupervised clustering of response related data. Then it attempts to relate the results to the physical, clinical and genetic attributes of the study population.

To evaluate the effectiveness and utility of ML techniques in analyzing patient response to the intervention vs. ‘typical’ responders analysis, this study used as a benchmark the Objective Response Overall Response (OR) over time, as defined by the study protocol [29]. The patient-subgroup characteristics of that were compared to those of subgroups defined by ML-based clustering methods, applied on raw and derived from raw outcome and response variables. For the purposes of benchmark analysis of response (referred to as OR) the response statuses can be summarized as: Partial Response (PR), Stable, Progressive Disease (PD) and Unable to Evaluate (UE) - no Complete Response status was recorded by the end of the study. To address limitations of sample size within the smallest status levels of the original variable, the OR status was analyzed as a binary variable (0 = PD and UE patients vs. 1 = PR and Stable patients). The benchmark OR approach of this study is consistent with the actual statistical analysis of responders conducted for the original study population.

As an alternative to benchmark OR-based approach, ML-based techniques are deployed to analyze outcome and response data collected. Unsupervised clustering analysis of outcome and response data was conducted, considering all available efficacy endpoint variables, including OR and haematology values change over time. Raw primary and secondary efficacy data related to time-series lesion measurements were engineered for the creation of new, tumour type-specific response variables. In total, 10 new variables were created based on

- the *absolute size and proportion of change (vs. baseline) in size over time* for every lesion type,
- the *timing of a tumour size change event that may indicate disease repression or progression or appearance event (new lesion)* and
- the *inter-individual range of response* based on target-lesion total size change compared to baseline measurement.

This feature engineering was to amplify the definition of response and hence capture the multivariate nature of response to treatment in terms of timing of response (time to response, duration of response and time to disease progression), magnitude (extent and range of response) and directionality (either positive or negative) for all lesion types assessed over time and available in the accessed dataset. ML-based multivariate unsupervised clustering was performed on the super-set of these derived response variables after pre-processing. Two approaches were examined within that: “Extended response” uses all aforementioned engineered response variables (10 in total), whilst “haematology enriched” approach builds on top of “extended response” to also include the changes in blood markers that are correlated to tumour burden based on existing knowledge on disease progression and overall survival predicting factors for CRC. Available biomarkers used to shed light on tumour burden and respective literature evidence on their association with CRC survival metrics were: Carcinoembryonic Antigen (CEA) [30], Lactate dehydrogenase (LDH) [31] and Alkaline Phosphatase (ALP) [32]. A summary view of the three approaches used to define response status and the data types included can be viewed in 2.1. The main hypothesis for the comparison of results across the three approaches is that a more comprehensive definition of response (extended response-based approaches) when analyzed by ML techniques can maximally dissect the differences in outcomes, endpoint values and timing of potential response events vs. that achieved with the benchmark approach within the same patient population. Furthermore, the partition of the study population based on statistically supported response heterogeneity could consequently correspond to optimally refined subgroups of patients with qualitative differences in physical, clinical and genetic characteristics.

### 2.2 Study Population

The trial enroled 463 mCRC patients, randomly assigned to receive Panitumumab plus BSC or BSC alone as a third- or fourth-line therapy. The main objective of that study was to determine whether the intervention improves Progression-Free Survival (PFS) time compared with BSC alone, followed by best Objective Response by blinded central review and Overall Survival (OS) time [28]. The study dataset was accessed via Datasphere [33], where only 80% of the RCT original data was available, whilst Objective Response, as defined by the study protocol [29], was recorded for 88% of the accessed patients. Subsequently, the patient population for this study, after exclusions, is 326 observations with a 35-week median follow-up time

### 2.3 Data pre-processing

All numeric variables were examined for outlying values and one patient was excluded at the basis of their PFS day being higher than 5 Standard Deviations from the mean. Missing data analysis on lesion assessment raw data revealed 30-37% missing. Given the commonalities in missing patterns across lesion types and the available OR values from the original dataset for those observations where lesion measurements were missing, it was assumed that lesion data was missing at random, due to disease progression or clinical state deterioration. Missing data were imputed with multiple imputation technique using the *MICE* package in R [34]. All numerical variables were used as predictors for the missing data variables values and Predictive Mean Matching (PMM) method was applied. The pooled results from 50 imputed datasets were used. Continuous numeric variables were normalized and scaled to a mean of 0 and a SD of 1 before applying clustering algorithms.

### 2.4 Descriptive Analysis

The first set of analyses aimed in exploring the data for signs and characteristics of response heterogeneity within each treatment arm and in the full sample. Various data exploration and visualization techniques, descriptive analysis, usual responders analysis and primary outcome prediction modeling were implemented. By exploring the distribution of baseline characteristics and outcome variables for each treatment arm and the full study population, we assessed the prognostic nature of baseline features like KRAS codon 12/13 and Eastern Cooperative Oncology Group (ECOG) status. Mixed models were applied to explore signs of differential response within the control group and on the full sample aiming to confirm the validity of feature engineering. Comparison of responders rates within the two treatment groups and sub-groups within those defined by prognostic factors, was performed with Chi-square and Cochran Mantel-Haenzsel tests and was based on Objective and derived response metrics. The true effect of the treatment to the response heterogeneity as measured by the change in target-lesions total size was calculated as:

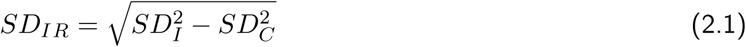

where *SD*_*IR*_ is the SD of the net individual response to Panitumumab and *SD*_*I*_ and *SD*_*C*_ are the SDs of the change in tumour size for the Intervention (Panitumumab) and Control (BSC) arms respectively [11]. PFS times for the two treatment groups were compared via Kaplan-Meier curves. They were also modeled against all demographic, clinical, genetic and endpoint related metrics via Cox proportional hazards regression analysis, adjusted for inter-individual variability, to illuminate potential confounders and effect modifiers with regards to response to treatment. Lastly, the similarity of the study population characteristics when grouped into responders and non-responders per the benchmark approach (OR-based) was studied via statistical tests.

### 2.5 ML-based Unsupervised Clustering Analysis

The purpose of applying ML clustering techniques on the extended response datasets is to detect hidden structure in the data as a means of unfolding and thus describing heterogeneity in treatment response for mCRC patient population. For this purpose, the study compares the performance of three most popular approaches of unsupervised clustering: distance-based Hierarchical Agglomerative Clustering (HAC), heuristic method of Partition Around Medoids (PAM) and model-based Gaussian Mixture Modelling (GMM) method. This allowed us to examine different scenarios of cluster relationship and data structure, in specific, hierarchical vs. overlapping vs. parametric and probabilistic methods to define belonging of a single patient data into distinct response profiles (clusters). PAM method was selected over fuzzy K-means clustering due to its superiority in outlier handling.

1. HAC: Compactness and separation of clusters across the different cut-off candidates was then surveyed with the Elbow and Silhouette plots.
2. PAM: The Gower’s distance was used to define the dissimilarity between data points and the size of clusters was determined also via the Elbow and Silhouette methods. Three medoids were configured to allow the algorithm start iterating across different sets of medoids and the optimal results were reached after 69 iterations.
3. GMM: Choosing the right number of clusters was addressed by statistical modeling and comparison of models with differing component distributions [35] in terms of Bayesian Information (BIC) score [36] and as evidence of fit for the corresponding model.

### 2.6 Validity, Stability and Sensitivity Analyses

The quality of clustering methods outputs was tested with multiple established *internal validity* and *stability* measures. This study used some of the most established cluster validity indexes, namely: Dunn’s, Silhouette and Connectivity index. The BIC index [36] was used only for the Gaussian mixture model-based clustering method. Stability analysis was used to test the “robustness of the clusters” or how the clusters resist being dissolved under plausible variations in the dataset. Examined stability measures included the average proportion of non-overlap (APN), the average distance (AD) and the figure of merit (FOM) [37]. Those metrics were used for relative validation as well, comparing their values across a range of choices for the optimal number of clusters (from 2 to 7) per clustering method. An alternative stability assessment approach was also performed via the package *clusterboot* in R [38] allowing for the calculation of the Jaccard index [39]. The data was sub-sampled 100 times and the Jaccard coefficient was calculated across the three applied clustering methods. The mean value was then used to assess cluster stability for each cluster in the original clustering of each method. The cluster-membership defined subgroup were described and compared in richness and strength of differentiation across the outputs of the three methods. The objective of such analysis was to discover the extent to which patients are qualitatively different from cluster to cluster and compare this degree and utility of differentiation vs. that provided by the benchmark approach, being the typical method for responders analysis in an RCT. In the absence of ground truth in sub-typing patients’ response to the intervention, this provides a way for measuring the correctness and relevance of the clustering and assists corroboration of existing evidence and formulation of hypothesis for deciphering further the response heterogeneity based on the biological characteristics present in the specific dataset. Sensitivity analysis was finally run using the same unsupervised clustering and validation methods onto the intervention treatment arm only (N=160 patients).

## 3 Results

### 3.1 Descriptive & “Traditional” Responders Analyses

The baseline characteristics of this study population (see Supplementary Data) are very similar to these of the original RCT data [28].

In total, 160 patients were treated with Panitumumab plus BSC and 166 with BSC alone. Based on Objective Response (OR) as defined by the study design, 92.2% of the BSC group patients were evaluated with ‘Progression of Disease’ or ‘Unable to Evaluate’ status after a minimum follow-up period of 52 weeks. This is statistically significant higher to 88.1% status concentration observed in the Panitumumab plus BSC group. The generalized Cochran-Mantel-Haenszel tests adjusted for the prognostic factors of ECOG and KRAS codon 12/13 status also provide strong evidence of difference between the two groups (p-values < and <.001 respectively). Overall, no patients achieved Complete Response and only 8 (2.5%) achieved Partial Response status, all of which were receivers of Panitumumab plus BSC treatment. Another 14.7% of total patients, almost equally split in the 2 treatment groups, was Stable by the end of the follow-up period whilst disease stabilization is proven to be clinically meaningful for mCRC patients [40]. The full results of the descriptive analysis are available in Supplementary Data. Key findings include:

- Mean PFS time, as the primary efficacy endpoint across the two treatment arms, when stratified by prognostic factor of KRAS exon 2 c12/13, is significantly different for the experimental vs. BSC alone arm.
- Comparison of responders rate based on target-lesions total size across the two treatment groups, adjusted for ECOG and KRAS c12/13 status provides additional evidence of response heterogeneity.
- The inter-individual difference in magnitude of response (*SD*_*IR*_ metric) due to treatment effect is larger than that attributed to random within-subject variability over time and has a clinically important effect size (+3.15cm).
- Days to disease progression or death were statistically significant improved for the Panitumumab plus BSC group compared with the BSC alone group (p-value <.001, stratified log-rank test). The difference in median PFS time between the two groups is almost doubled when further stratifying the populations for KRAS c12/13 status.
- Patients who received the experimental treatment had almost half the disease progression hazard compared to that of the BSC alone treatment group (HR: 0.59, 95%CI from 0.46 to 0.76). Longer time since disease diagnosis at baseline results in improved PFS times when adjusted for all other risk and prognostic factors, possibly suggesting differential mechanisms of resistance to disease.
- Overall, the two “responders” groups as defined by OR status differ significantly in only 2 out of 19 total baseline characteristics and in 12 out of 20 examined outcome and response characteristics (Table 3.1).

**Table 3.1:** Baseline, outcome & response characteristics by Objective Response OR (Benchmark Approach)

| Characteristic | "Non-responders" group (PD or UE) (N=270) | "Responders" group (Stable or PR) (N=56) | P-value |
| --- | --- | --- | --- |
| Age | 60.86 (10.06) | 61.41 (9.70) | 0.7 |
| Sex |  |  | 0.554 |
| Female | 101 (37.4) | 18 (32.1) |  |
| Male | 169 (62.6) | 38 (67.9) |  |
| Had previous surgery |  |  | 1 |
| No | 7 (2.6) | 1 (1.8) |  |
| Yes | 263 (97.4) | 55 (98.2) |  |
| Months since diagnosis† | 30.28 (17.86) | 37.69 (20.75) | 0.015 |
| Baseline Weight (kg) | 74.26 (15.74) | 74.49 (15.33) | 0.92 |
| Baseline ECOG status |  |  | 0.273 |
| Fully active | 111 (41.1) | 28 (50.0) |  |
| Symptoms but ambulatory | 133 (49.3) | 21 (37.5) |  |
| In bed <50% of the time | 26 (9.6) | 7 (12.5) |  |
| Histological Subtype |  |  | 0.365 |
| Mucinous | 26 (9.6) | 9 (16.1) |  |
| Other | 30 (11.1) | 6 (10.7) |  |
| No Subtype | 214 (79.3) | 41 (73.2) |  |
| Primary diagnosis |  |  | 0.978 |
| Colon | 177 (65.6) | 36 (64.3) |  |
| Rectal | 93 (34.4) | 20 (35.7) |  |
| Biomarker KRAS exon 2(c12/13) |  |  | 0.194 |
| Mutant | 95 (35.2) | 27 (48.2) |  |
| Wild-type | 154 (57.0) | 25 (44.6) |  |
| Failure | 21 (7.8) | 4 (7.1) |  |
| Baseline CEA (ng/ml) | 728.16 (1659.77) | 480.54 (1103.51) | 0.169 |
| Baseline LDH (U/L) | 502.54 (527.35) | 423.46 (520.17) | 0.305 |
| Baseline ALP (U/L) | 235.80 (229.29) | 204.93 (162.94) | 0.235 |
| Baseline Albumin (g/L) | 39.25 (4.47) | 39.80 (5.23) | 0.462 |
| Baseline Creatinine (umol/L) | 70.06 (21.03) | 70.84 (22.00) | 0.808 |
| Baseline Haemoglobin (g/L) | 124.88 (17.03) | 127.93 (17.40) | 0.234 |
| Baseline Platelets (10<sup>9</sup>/L)† | 268.71 (105.77) | 230.07 (86.67) | 0.004 |
| Baseline WBC (10<sup>9</sup>/L) | 8.37 (3.26) | 7.77 (2.47) | 0.127 |
| Baseline non-target lesions (units) | 6.07 (2.64) | 5.79 (2.53) | 0.443 |
| Baseline target-lesions size (mm) | 194.10 (94.69) | 185.43 (74.40) | 0.452 |
| Treatment |  |  | 0.143 |
| BSC alone | 132 (48.9) | 34 (60.7) |  |
| Panitumumab plus BSC | 138 (51.1) | 22 (39.3) |  |
| Weight change* | -0.83 (3.26) | -1.14 (2.93) | 0.481 |
| CEA change* | 288.68 (1164.62) | 183.57 (746.12) | 0.392 |
| LDH change*† | 196.49 (544.45) | 41.82 (212.78) | <0.001 |
| ALP change* | 83.54 (191.16) | 39.92 (146.40) | 0.058 |
| Days to death event*† | 248.52 (182.11) | 319.18 (218.61) | 0.027 |
| Death event* |  |  | 0.066 |
| No | 20 (7.4) | 9 (16.1) |  |
| Yes | 250 (92.6) | 47 (83.9) |  |
| Progression Free Survival time† | 70.97 (67.07) | 170.07 (140.21) | <0.001 |
| Disease progression during study† |  |  | <0.001 |
| No | 3 (1.1) | 12 (21.4) |  |
| Yes | 267 (98.9) | 44 (78.6) |  |
| Objective Response: Target-lesions size† |  |  | <0.001 |
| PD | 161 (59.6) | 1 (1.8) |  |
| PR | 2 (0.7) | 8 (14.3) |  |
| Stable | 86 (31.9) | 39 (69.6) |  |
| UE | 21 (7.8) | 8 (14.3) |  |
| Time to PD by target-lesions size | 111.00 (91.08) | 128.43 (108.53) | 0.265 |
| Target-lesions size change (mm)*† | 45.22 (34.64) | 9.87 (34.66) | <0.001 |
| Target-lesions size change rate*† | 0.24 (0.19) | 0.04 (0.16) | <0.001 |
| Target-lesions max size (mm)*† | 245.91 (115.37) | 211.85 (84.27) | 0.012 |
| Target-lesions max increase (mm)*† | 50.84 (32.01) | 24.28 (20.75) | <0.001 |
| Target-lesions min size (mm)* | 187.33 (95.25) | 170.44 (70.72) | 0.131 |
| Target-lesions min size day | 12.30 (42.80) | 24.02 (44.61) | 0.076 |
| Ind. range of response (target-lesions size)*† | -40.05 (38.83) | -7.34 (40.74) | <0.001 |
| New lesion(s) event* |  |  | 1 |
| No | 16 (5.9) | 3 (5.4) |  |
| Yes | 254 (94.1) | 53 (94.6) |  |
| New lesion(s) event day† | 79.57 (59.83) | 172.27 (120.29) | <0.001 |
| New lesions size last visit (mm)† | 23.94 (18.52) | 16.87 (8.21) | <0.001 |

Counts (percentages) used for categorical variables, means (SDs) for continuous numerical variables. Qualitative differences across characteristics were tested with chi-square for categorical variables and regular ANOVA one-way tests for continuous variables. A P-value of <0.05 was considered statistically significant.
\*recorded during the study and up to the end of follow-up period
† statistically significant difference

OR-based responders are diseased for a statistically significant longer period of time at baseline vs. nonresponders (37.7 vs. 30.3 months, p-value=0.015) possibly suggesting inherent disease resistance behaviour.

Allocation to treatment is balanced for the two groups and therefore does not associate with differential response. Significant differences are observed in primary outcome metrics and most derived response measures (based on target-lesions size and new lesion events) are also significantly different between the two groups proving their utility as proxy or superior to OR measures.

### 3.2 Clustering of Study Population based on Response

Three different ML-based techniques were used on the extended response and the further augmented with haematology data extended response datasets. Hierarchical Agglomerative Clustering on extended response dataset and the configuration of 3 clusters (clusters size: 240, 39 and 47 patients) was chosen based on internal quality measures. A t-SNE visualization of the data indicates a clear visual separation of the 3 clusters based on composite response patterns. Partitioning the same data with the PAM algorithm suggested the formation of 7 distinct clusters (clusters size: 82, 29, 33, 32, 74, 16 and 61 patients) based on internal quality measures. Despite the increase in partition, the separation of the 7 clusters remains neat when visualized with t-SNE. Lastly, finite mixture model-based clustering technique and the configuration of five clusters as chosen based on the BIC criterion formed of groups of varying volume, ellipsoidal shape and varying orientation (clusters size: 58, 31, 75, 70 and 92 patients). Visual inspection of the 5 groups on a two-dimensional space shows a fair amount of overlap across the groups. The two-dimensional plot visualizations of the three algorithms output can be viewed in Fig 3.1. The same three algorithms and quality appraisal measures were applied onto the haematology data augmented outcome & response metrics dataset, attempting to evaluate the ability of the algorithms to refine partitioning of the population based also on the change in blood markers, as a proxy to tumour burden. Both HAC and PAM clustering methods yield best quality results when configured for two only clusters. The partitioning based on HAC results into clusters of very similar size and structure with the ones obtained by the OR-based approach. Partitioning by PAM method suggests a more balanced divide whilst GMM proposed three clusters and therefore provided a finer separation of the population.

**Figure 2.1:**
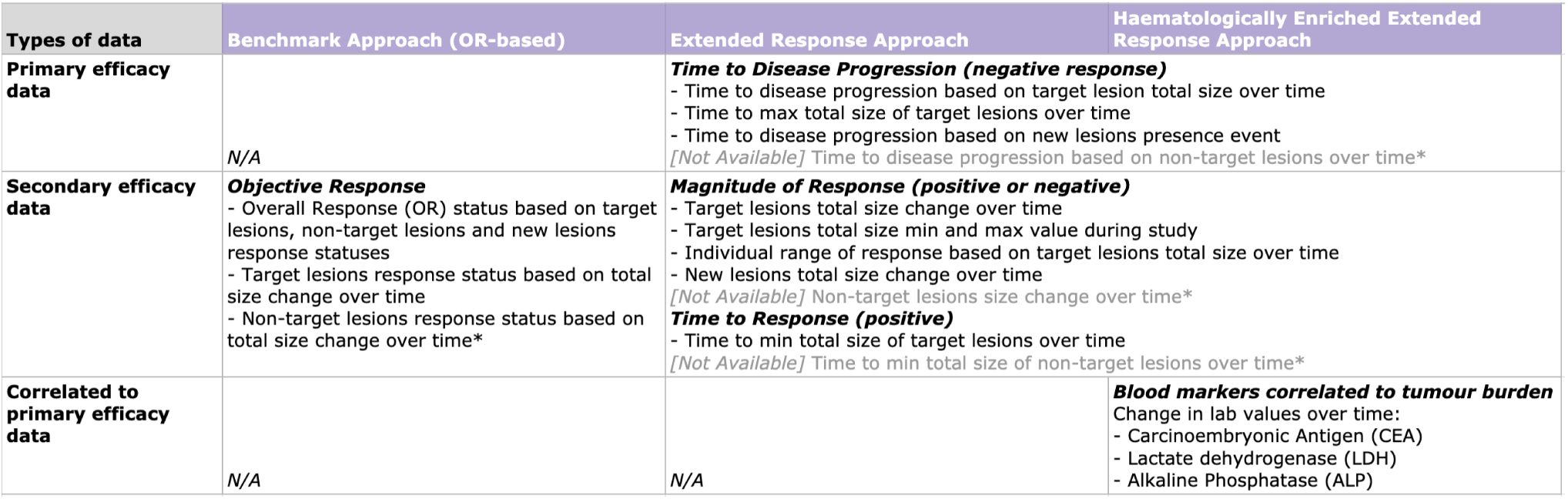
Response definition framework: Approaches & Data types. *Raw measurements for non-target lesions size were not available in the dataset and therefore non-target lesions size change data has only been included for OR status determination (original dataset variable) in Benchmark Approach.

**Figure 3.2:**
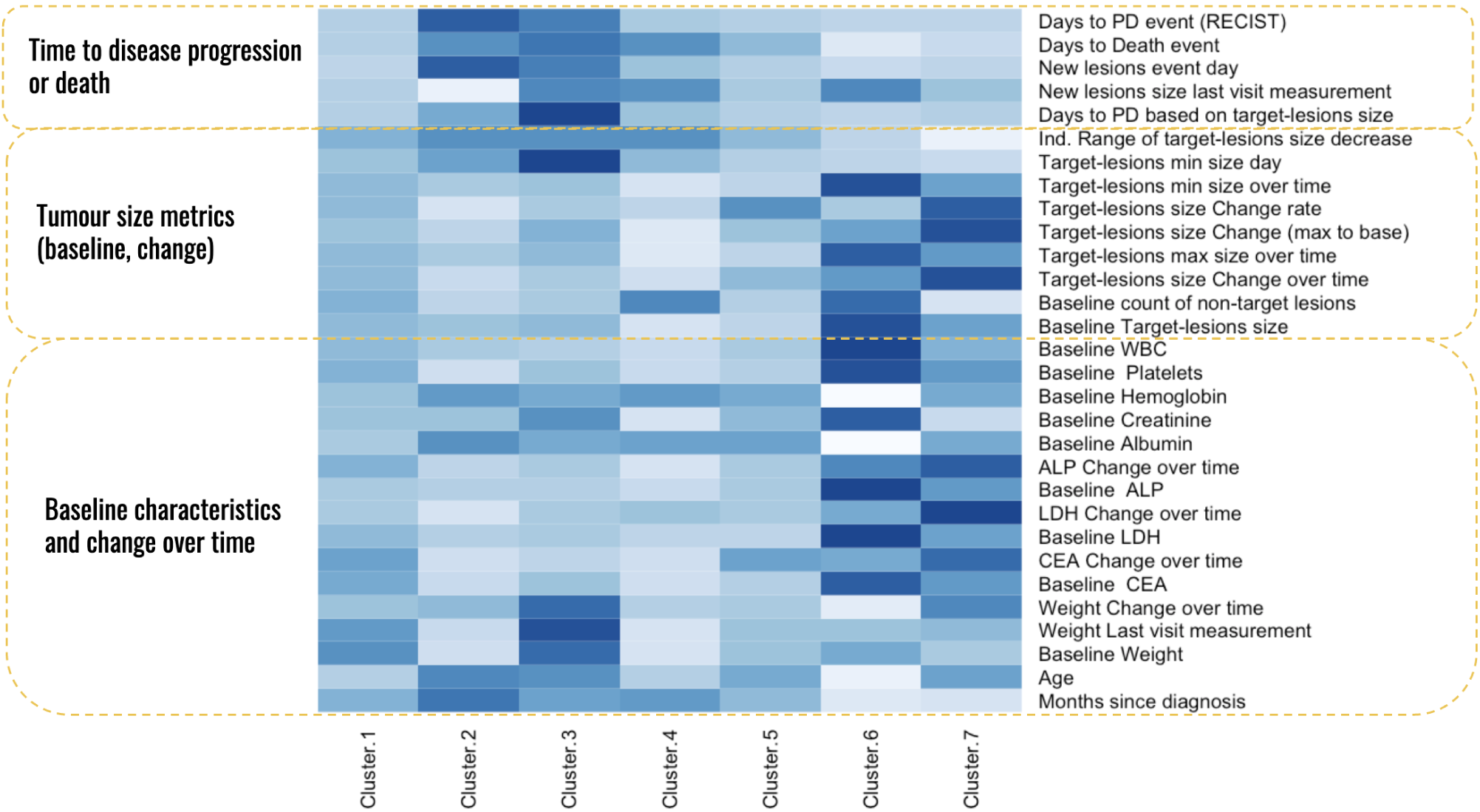
Heatmap visualization of differentiation within PAM-based clusters across all baseline characteristics, over time outcome and response metrics.

### 3.3 Internal Validation and Stability Analysis of Clustering Results

The goodness of the clustering methods was evaluated by various internal validity measures and compared across approaches (see Supplementary Data). Based on internal validity metrics HAC and PAM methods applied on the extended response dataset, proposed clusters of similar quality, proven by maximal compactness and separability values, when compared to the same metrics for model-based clustering algorithm (see Supplementary Data). Addition of CEA, LDH and ALP markers changes over time to the extended response data seems to introduce noise for the algorithm and thus, cross-cluster differences in outcome and response metrics are attenuated.

PAM’s partitioning of the study population into 7 clusters outperformed all other clustering methods (see Supplementary Data) with the lowest Jaccard coefficient recorded being 0.68 (cluster 3). Two clusters revealed by HAC method had only 0.37 Jaccard’s coefficient, indicating highly unstable groupings. GMM’s clusters were equally unstable. Other examined stability measures results were consistent with these findings.

### 3.4 External Validation of Clustering Results

The last set of analyses performed on the extended response dataset was an in-depth comparison of the multi- variate nature characteristics of the study population on the basis of their cluster membership. As per above, PAM method suggests the most extensive and robust partitioning of observations based on outcome and response data. Figure 3.2 provides a visual representation of the relative degree of differentiation within the 7 clusters across all examined outcome, response and baseline variables. Highest values in outcome metrics suggesting response or resistance to disease progression are concentrated in clusters 2 and 3. Conversely, highest change values over time across most of the response metrics, including blood markers correlated to tumour burden, are recorded in clusters 6 and 7, potentially suggesting increased heterogeneity in response metrics as also observed during exploratory analysis of sub-populations. The effect size of the differentiation observed within the distinct groups defined by PAM membership can be viewed in Table 3.2. Overall, the seven subgroups differ significantly in 8 out of 19 baseline characteristics and in 17 out of 20 examined outcome and response characteristics. Evidence of association between the treatment type and differential outcomes is also provided for the seven subgroups. Compared to the sub-typing inferred by the benchmark approach, PAM cluster membership results into more subgroups and greater differentiation within those across most available characteristics.

**Table 3.2:** Baseline, outcome and response characteristics of patient subgroups based on cluster membership (PAM)

| Characteristic | Cluster 1 | Cluster 2 | Cluster 3 | Cluster 4 | Cluster 5 | Cluster 6 | Cluster 7 |
| --- | --- | --- | --- | --- | --- | --- | --- |
| Age | 60.2 (9.1) | 62.1 (11.0) | 61.8 (11.2) | 60.1 (10.9) | 61.3 (9.5) | 58.8 (11.4) | 61.5 (10.0) |
| Sex |  |  |  |  |  |  |  |
| Female | 29 (35.4) | 11 ( 37.9) | 10 ( 30.3) | 14 ( 43.8) | 33 ( 44.6) | 2 ( 13.3) | 20 ( 32.8) |
| Male | 53 (64.6) | 18 ( 62.1) | 23 ( 69.7) | 18 ( 56.2) | 41 ( 55.4) | 13 ( 86.7) | 41 ( 67.2) |
| Had previous surgery |  |  |  |  |  |  |  |
| No | 2 ( 2.4) | 1 ( 3.4) | 0 ( 0.0) | 0 ( 0.0) | 1 ( 1.4) | 0 ( 0.0) | 4 ( 6.6) |
| Yes | 80 (97.6) | 28 ( 96.6) | 33 (100.0) | 32 (100.0) | 73 (98.6) | 15 (100.0) | 57 ( 93.4) |
| Months since diagnosis† | 32.4 (19.9) | 40.1 (21.8) | 35.4 (16.7) | 36.43 (15.64) | 31.6 (18.4) | 23.4 (14.6) | 23.7 (15.5) |
| Baseline Weight (kg) | 75.6 (17.2) | 72.4 (14.9) | 76.8 (15.3) | 71.9 (12.2) | 73.9 (15.7) | 74.8 (15.1) | 73.6 (15.9) |
| Baseline ECOG status |  |  |  |  |  |  |  |
| Fully active | 30 (36.6) | 16 ( 55.2) | 20 ( 60.6) | 15 ( 46.9) | 30 (40.5) | 5 ( 33.3) | 23 ( 37.7) |
| Symptoms but ambulatory | 40 (48.8) | 10 ( 34.5) | 12 ( 36.4) | 15 ( 46.9) | 40 (54.1) | 6 ( 40.0) | 31 ( 50.8) |
| In bed < 50% of the time | 12 (14.6) | 3 ( 10.3) | 1 ( 3.0) | 2 ( 6.2) | 4 ( 5.4) | 4 ( 26.7) | 7 ( 11.5) |
| Histological Subtype |  |  |  |  |  |  |  |
| Mucinous | 8 ( 9.8) | 5 ( 17.2) | 1 ( 3.0) | 6 ( 18.8) | 5 ( 6.8) | 1 ( 6.7) | 9 ( 14.8) |
| Other | 10 (12.2) | 2 ( 6.9) | 7 ( 21.2) | 4 ( 12.5) | 8 (10.8) | 1 ( 6.7) | 4 ( 6.6) |
| No Subtype | 64 (78.0) | 22 ( 75.9) | 25 ( 75.8) | 22 ( 68.8) | 61 (82.4) | 13 ( 86.7) | 48 ( 78.7) |
| Primary diagnosis |  |  |  |  |  |  |  |
| Colon | 52 (63.4) | 17 ( 58.6) | 19 ( 57.6) | 23 ( 71.9) | 51 (68.9) | 10 ( 66.7) | 41 ( 67.2) |
| Rectal | 30 (36.6) | 12 ( 41.4) | 14 ( 42.4) | 9 ( 28.1) | 23 (31.1) | 5 ( 33.3) | 20 ( 32.8) |
| Biomarker KRAS (c12/13) |  |  |  |  |  |  |  |
| Mutant | 37 (45.1) | 9 ( 31.0) | 4 ( 12.1) | 15 ( 46.9) | 30 (40.5) | 5 ( 33.3) | 22 ( 36.1) |
| Wild-type | 43 (52.4) | 16 ( 55.2) | 25 ( 75.8) | 14 ( 43.8) | 39 (52.7) | 8 ( 53.3) | 34 ( 55.7) |
| Failure | 2 ( 2.4) | 4 ( 13.8) | 4 ( 12.1) | 3 ( 9.4) | 5 ( 6.8) | 2 ( 13.3) | 5 ( 8.2) |
| Treatment† |  |  |  |  |  |  |  |
| BSC alone | 47 (57.3) | 15 ( 51.7) | 1 ( 3.0) | 19 ( 59.4) | 42 (56.8) | 8 ( 53.3) | 34 ( 55.7) |
| Panitumumab plus BSC | 35 (42.7) | 14 ( 48.3) | 32 ( 97.0) | 13 ( 40.6) | 32 (43.2) | 7 ( 46.7) | 27 ( 44.3) |
| Baseline CEA (ng/ml)† | 831.9 (1507.7) | 250.0 (363.9) | 631.5 (1010.9) | 180.3 (353.9) | 482.9 (861.0) | 1567.8 (1777.9) | 1019.4 (2741.7) |
| Baseline LDH (U/L)† | 516.1 (504.2) | 353.2 (469.3) | 413.9 (308.8) | 292.7 (311.2) | 329.9 (218.9) | 1193.8 (1035.4) | 680.2 (646.8) |
| Baseline ALP (U/L) | 220.6 (175.0) | 210.3 (145.2) | 202.2 (137.8) | 184.3 (275.4) | 215.3 (196.7) | 354.8 (256.8) | 280.9 (298.6) |
| Baseline Albumin (g/L)† | 38.2 (4.9) | 40.7 (5.1) | 39.8 (3.1) | 40.1 (4.4) | 40.1 (4.3) | 35.1 (4.9) | 39.7 (4.3) |
| Baseline Creatinine (umol/L) | 70.0 (22.3) | 70.0 (22.0) | 74.3 (22.6) | 66.3 (17.0) | 70.8 (22.1) | 77.3 (27.8) | 67.9 (17.3) |
| Baseline Haemoglobin (g/L)† | 123.0 (15.7) | 129.5 (17.7) | 127.0 (14.3) | 129.1 (15.2) | 126.8 (19.6) | 107.5 (15.7) | 126.7 (15.6) |
| Baseline Platelets (10<sup>9</sup>/L)y | 269.5 (101.9) | 228.2 (80.9) | 256.2 (117.2) | 236.9 (106.2) | 249.1 (92.4) | 323.6 (148.4) | 285.1 (98.8) |
| Baseline WBC (10<sup>9</sup>/L)y | 8.4 (2.8) | 8.0 (2.0) | 7.8 (3.1) | 7.0 (2.4) | 7.8 (2.5) | 11.7 (6.5) | 8.8 (3.2) |
| Baseline target-lesions size (mm)† | 208.1 (37.3) | 177.9 (57.6) | 210.3 (51.5) | 62.6 (26.2) | 131.0 (50.7) | 417.5 (70.6) | 256.9 (68.6) |
| Baseline non-target lesions (units)† | 6.3 (2.5) | 5.7 (2.8) | 6.0 (1.6) | 7.0 (2.8) | 5.8 (2.5) | 7.5 (3.2) | 5.2 (2.7) |
| Weight change* | -1.1 (3.7) | -0.9 (2.5) | 0.1 (4.2) | -1.4 (2.9) | -1.2 (2.6) | -2.0 (3.0) | -0.3 (2.8) |
| CEA change* | 309.1 (1426.4) | 68.2 (452.7) | 109.2 (1578.5) | 72.4 (238.1) | 302.2 (859.0) | 275.9 (856.3) | 466.9 (1110.2) |
| LDH change*† | 114.5 (467.9) | 8.1 (202.1) | 109.0 (473.8) | 143.5 (612.5) | 123.1 (233.5) | 200.5 (764.4) | 417.5 (688.2) |
| ALP change*† | 84.3 (156.9) | 25.1 (129.3) | 46.1 (168.2) | -18.6 (109.5) | 40.1 (103.8) | 137.2 (199.7) | 183.5 (281.7) |
| Objective Response OR† |  |  |  |  |  |  |  |
| PD or UE | 81 (98.8) | 2 ( 6.9) | 32 ( 97.0) | 31 ( 96.9) | 74 (100.0) | 13 ( 86.7) | 61 (100.0) |
| PR or Stable | 1 ( 1.2) | 27 ( 93.1) | 1 ( 3.0) | 1 ( 3.1) | 0 ( 0.0) | 2 ( 13.3) | 0 ( 0.0) |
| Objective Response (Target-lesions)† |  |  |  |  |  |  |  |
| PD or UE | 19 (23.2) | 3 ( 10.3) | 28 ( 84.8) | 5 ( 15.6) | 74 ( 100.0) | 1 ( 6.7) | 61 (100.0) |
| PR or Stable | 63 (76.8) | 26 ( 89.7) | 5 ( 15.1) | 27 ( 84.3) | 0 ( 0.0) | 14 ( 93.3) | 0 ( 0.0) |
| Target-lesions size change (mm)*† | 32.4 (15.2) | 6.1 (40.8) | 25.4 (30.7) | 4.8 (11.8) | 33.4 (17.3) | 55.9 (30.1) | 92.25 (30.12) |
| Target-lesions max size (mm)*† | 249.8 (39.1) | 207.7 (70.2) | 259.6 (48.6) | 70.0 (29.1) | 169.3 (60.5) | 473.5 (92.8) | 349.4 (79.5) |
| Target-lesions max increase (mm)*† | 40.6 (15.8) | 26.6 (21.5) | 45.4 (16.8) | 7.4 (7.6) | 37.3 (17.6) | 55.9 (30.1) | 92.6 (29.4) |
| Target-lesions size change rate† | 0.2 (0.1) | 0.0 (0.2) | 0.1 (0.2) | 0.1 (0.2) | 0.3 (0.2) | 0.1 (0.1) | 0.4 (0.2) |
| Target-lesions min size (mm)*† | 200.5 (39.0) | 159.3 (51.5) | 177.9 (47.6) | 59.7 (26.0) | 126.6 (48.2) | 417.5 (70.6) | 256.6 (68.3) |
| Target-lesions min size day† | 14.4 (28.9) | 34.9 (53.2) | 84.6 (40.0) | 17.2 (55.9) | -1.4 (28.1) | -6.8 (7.1) | -10.8 (16.1) |
| Ind. range target-lesions response*† | -27.3 (14.4) | -5.7 (41.0) | -2.0 (47.7) | -4.4 (12.1) | -30.3 (22.8) | -55.9 (30.1) | -90.7 (32.0) |
| Days to PD (target-lesions based)† | 83.0 (62.3) | 154.0 (119.1) | 298.9 (96.1) | 114.4 (98.0) | 86.5 (39.0) | 74.7 (40.7) | 79.28 (28.7) |
| New lesion(s) event* |  |  |  |  |  |  |  |
| No | 5 ( 6.1) | 1 ( 3.4) | 2 ( 6.1) | 2 ( 6.2) | 3 ( 4.1) | 0 ( 0.0) | 6 ( 9.8) |
| Yes | 77 (93.9) | 28 ( 96.6) | 31 ( 93.9) | 30 ( 93.8) | 71 (95.9) | 15 (100.0) | 55 ( 90.2) |
| New lesions size last visit (mm)† | 20.8 (13.3) | 14.8 (7.3) | 28.8 (20.1) | 27.9 (24.8) | 22.0 (16.9) | 28.7 (15.6) | 22.51 (18.87) |
| New lesion(s) event day† | 60.6 (33.0) | 240.1 (118.9) | 209.3 (54.0) | 96.6 (68.2) | 69.3 (33.6) | 45.8 (23.8) | 55.4 (20.45) |
| Days to Death event† | 200.5 (133.5) | 376.9 (202.6) | 443.8 (177.0) | 383.2 (244.4) | 270.8 (169.8) | 101.4 (80.1) | 149.7 (109.9) |
| Death event* |  |  |  |  |  |  |  |
| No | 1 ( 1.2) | 6 ( 20.7) | 5 ( 15.2) | 6 ( 18.8) | 4 ( 5.4) | 1 ( 6.7) | 6 ( 9.8) |
| Yes | 81 (98.8) | 23 ( 79.3) | 28 ( 84.8) | 26 ( 81.2) | 70 (94.6) | 14 ( 93.3) | 55 ( 90.2) |
| Progression Free Survival time† | 53.2 (33.8) | 245.8 (136.6) | 205.4 (102.4) | 80.6 (86.7) | 56.1 (29.1) | 44.9 (24.5) | 49.4 (18.7) |
| Disease Progression event*† |  |  |  |  |  |  |  |
| No | 5 ( 6.1) | 5 ( 17.2) | 0 ( 0.0) | 2 ( 6.2) | 1 ( 1.4) | 2 ( 13.3) | 0 ( 0.0) |
| Yes | 77 (93.9) | 24 ( 82.8) | 33 (100.0) | 30 ( 93.8) | 73 (98.6) | 13 ( 86.7) | 61 (100.0) |

Counts and percentages are used for categorical variables and means and standard deviations for continuous numerical variables. Qualitative differences across characteristics were tested with chi-square test for categorical variables and regular ANOVA one-way test for continuous variables. Exact test and non-normal tests were used where required. A P-value of <0.05 was considered statistically significant.
\*recorded during the study and up to the end of follow-up period
† statistically significant difference

**Figure 3.1:**
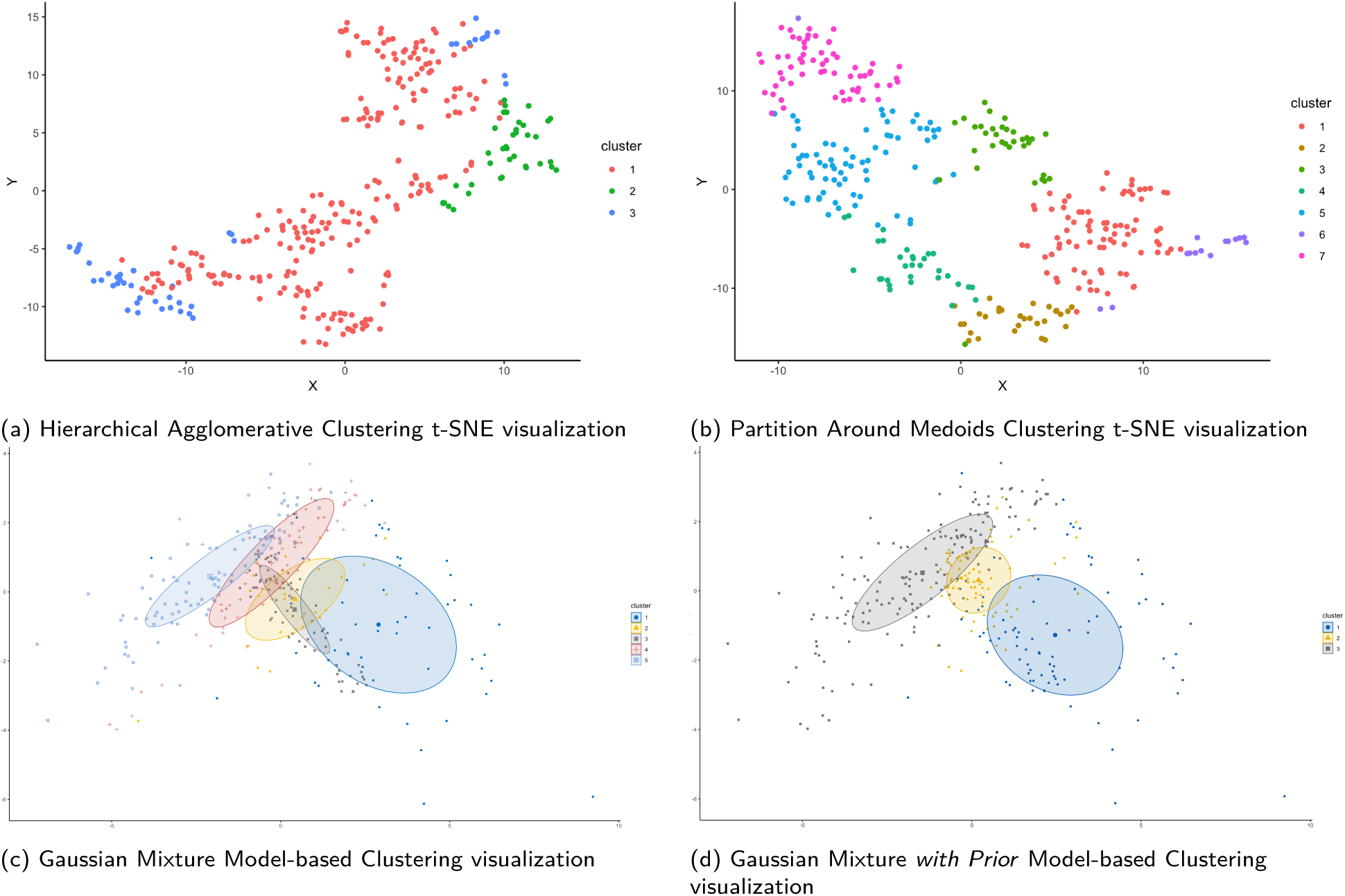
Extended Response dataset: Partition of study population based on outcome and response metrics via the use of ML-based clustering algorithms

### 3.5 Characterizing Patients based on Cluster Membership

Patients in clusters 2, 3, 4 and 6 have the highest percentages of PR or stable disease profiles, while clusters 1, 5 and 7 are made almost exclusively of patients with PD or UE status. A close inspection of the clusters representative of responders and non-responders groups reveals though that the divide obtained with PAM clustering justifies a differential clinical profile for each of the seven subgroups. As an example, Cluster 2, appears to be the *Disease Resistant* group. Overall its patients have better prognosis at baseline whilst being the longest diseased by the time of study entry. Although equally split into the two treatment arms, all patients manifest resistance to disease progression and achieve extended PFS and OS times. Their response profile is characterized by no change on target-lesions size (average change: +1%). There is evidence of tumour shrinkage during the first 5 weeks of the study whilst new lesions were relatively small and appeared late during the study follow-up period (month 8 on average). Cluster 3 can be characterized as the *True Positive Responders to Intervention* subgroup. Its patients have an average prognosis at baseline, whilst almost all (97%) received Panitumumab plus BSC treatment. Consistent with the evidence that the interventional treatment favours PFS times, this group achieved maximum response by magnitude and record-high PFS and OS times. Over 75% of this cluster members had KRAS exon 2 c12/13 wild-type status, an oncogene marker proven to respond positively to monoclonal antibody therapies [3]. Evidence shows that disease progression is presumably driven by the presence of new lesions which appeared on average on month 7 but progressed fast, possibly suggesting a new pattern of resistance to treatment.

Clusters 4, 6 and 7 are all *Negative Responders* subgroups and present poor outcomes and response metrics. However, magnitude of change in tumour sizes (target and new lesions), blood markers values change over time and speed of progression patterns still reveal differential profiles of response resistance.

### 3.6 Sensitivity Analysis

Partitioning of the Panitumumab plus BSC patients group only by PAM method resulted into three distinct clusters characterized by similar to the primary analysis quality and stability metrics. The null hypothesis of no difference between the characteristics of OR-driven and cluster membership-driven divide of patients into subgroups was also rejected in this case. ML-driven response identification revealed three groups representative of excellent, good and poor response, manifested by multiple response parameters and therapy outcomes and differentiated by KRAS exon 2 c12/13 status and incumbent metastatic activity (see Supplementary Data).

## 4 Discussion

### 4.1 Key Findings

This study demonstrates the utility of adopting ML-based techniques in identifying and characterizing responders of an RCT. Having tested multiple methods that overcome point limitations of the traditional responders analysis and contrasting the results of the clustering membership-based patient profiles with the former, we showed that traditional Objective Response (OR) based analysis is inferior in discovering heterogeneity in outcome and response and relating those to patient clinical and genetic characteristics.

Characterization of patients based on conventional analysis of response to the studied anti-EGFR drug resulted into ‘poor’ profiling information of the two subgroups, as “responders” and “non-responders” differ in a statistically significant and clinically relevant way only in 2 baseline characteristics. ML-based clustering of the study population based on outcome and response characteristics, as an alternative to OR-based responders identification offered much richer and granular findings on both the clinical and response profile of the study population. The seven statistically diverse subgroups suggested by PAM method clustering were differentiated in almost every examined response variable and characterized by different magnitude of tumour size change and disease progression patterns. As hypothesized, differentiation in outcome and response corresponds to qualitatively different baseline profiles across the seven groups. Improved survival times sub-populations are characterized by KRAS exon 2 c12/13 negative status and better prognosis at baseline (ECOG status and blood markers related to tumour burden). Conversely, over 50% of patients with KRAS negative status who were exposed to treatment experienced disease progression in early stages of the study, signifying heterogeneity in response even in populations with good genetic prognostic markers. Also, there is evidence of differential disease advancement behaviour for patients who did not receive the intervention.

Meta-analysis of anti-EGFR drugs effectiveness on mCRC patients provides clear evidence on response favouring KRAS exon 2 wild-type status [41, 42], however, further investigation of tumour mutational data detected mutations in KRAS, EGFR and several other genes as potential mechanisms of primary resistance to an antiEGFR mAb therapy [43, 44]. Even within wild-type KRAS exon 2 patients, other RAS mutations have been associated with poor response to Panitumumab [45]. The recorded statistically significant differences in blood markers associated with tumour burden across the seven subgroups support existing first-line treatment patients evidence [30, 31, 32, 46].

### 4.2 Strengths & Limitations

Although our study comes over 10 years after the evidence supporting effectiveness of anti-EGFR drug for mCRC patients and increased response in KRAS exon 2 c12/13 negative status populations, the application of the methodology used is timeless. The potential of getting to such rich information on factors predicting response or resistance to a therapy through re-analysing RCT data, even years after testing the effectiveness of a drug, may still affect clinical trial design/strategy and inform disease management in clinical practice. Additionally, the applied methods support leveraging domain expertise to engineer raw response data combined with ML techniques of reinforced interpretability. Such approaches are imperative towards more data-driven precision-medicine efforts as they allow the critiquing and auditing of decisions. Conversely, the utility of this study is limited by data-related and methodological factors. Generalization of findings across populations is limited by the predominantly white ethnicity of the studied RCT population. Findings were also limited by the data that was available as a secondary source vs. the data originally collected for the RCT. Furthermore, although overall stability of results was demonstrated, as with any statistical modelling, cautiousness is needed in guaranteeing generalizability to different input data. Finally, the study design within the RCT setting limits our ability to analyze the results in a counterfactual way, with no easy remedies to such limitations. Cross-over or multiple treatment designs adoption is neither a feasible nor an ethical alternative for such advanced stage metastatic patients, therefore, occasional variation of results at the patient level and their implications to overall findings cannot be excluded.

### 4.3 Clinical, Research Utility & Further Work

A plethora of resistance mechanisms and pathways related to inter-and intra-heterogeneity of tumour involving molecular and epigenetic abnormalities is available today to inform PPM practices. Nowadays that so much more data is being gathered around mCRC patients molecular and epigenetic characteristics, the potential of ML-based techniques application in identifying differential responses could be a game-changer for the efficacy and efficiency of future clinical trials. An ML-based exploration of response data in Phase II trials and a more refined characterization of responders in previous RCTs could pave the way to the design of drugs and patient selection that have significantly higher chances of success.

External validation of the approach via its application to another RCT dataset with similar baseline characteristics and cohort outcomes or to the extended, original dataset of this RCT could assure that similar informative and robust clustering can occur across datasets using the same methodology and that the pattern is not peculiar to the original dataset. Furthermore, extending the dataset to include optimal sequencing genetic, epigenetic, biomarker and quality of life data can equally extend the research question to phenotyping and validation of existing molecular sub-types of CRC based on the same response data.

### 4.4 Conclusions

We implemented Machine-Learning unsupervised clustering methods to analyze responses to antiEGFR drug among mCRC patients and to identify characteristics of non-responders. Partitioning by PAM method shows promise in uncovering heterogeneity in response and describing patterns across multiple endpoints over time, thus delineating hidden structure in data that is typically analyzed through composite measures like objective response in RCT practice.

We showed the efficiency of ML-based techniques in assessing the prognostic nature of collected baseline biomarkers, genetic and other clinical characteristics of the population by scrutinizing the differences in the distinct cluster-based cohorts.

Given that CRC heterogeneity is multi-faceted and diversity in response is manifested through multiple resistance mechanisms of primary and acquired resistance, further studies involving larger, more diverse and richer in genetic and clinical history information are warranted to corroborate the utility of the proposed methods in deciphering response to interventions. Nevertheless, this study is a ground-breaking attempt to change the paradigm of RCT responders analysis by showcasing the utility of mining existing RCT data to rationalize future clinical trials and increase their effectiveness in an accelerated pace.

## Data Availability

The data used for the conduct of this research is available through DataSphere portal.

https://www.datasphere.online/en/

## 5 Acknowledgements

The author thanks Paul Agapow for originating this research study and for the validity and actuality of its primary aim. His direction during literature review and his scientific work and deep expertise in applying Machine Learning techniques onto Oncology datasets have been invaluable for the definition of the methodological approach as well as for execution.

As this article is based on a research project in partial fulfillment of the MSc degree Health Data Analytics Machine Learning of Imperial College London, the authors also thank Imperial College and the Public School of Health in specific for their scientific and research staff support as well as access to high-computing resources.

