## Supplementary Data for "Using Machine-Learning Techniques to Identify Responders vs. Non-responders in Randomized Clinical Trials"

#### 1 Descriptive Analysis: Baseline characteristics of study population by treatment allocation

#### 2 Responders & Response Patterns Exploratory Analysis

Exploration of PFS time as the primary efficacy endpoint across the two treatment arms, when stratified by prognostic factors of ECOG and KRAS exon 2 c12/13 status, provides strong evidence of difference in mean days to disease progression for the experimental vs. BSC alone arm (mean PFS for KRAS negative patients within BSC alone group is 64.9 vs. 128.9 days for the Panitumumab plus BSC group; mean PFS for ECOG 1 or 2 baseline status within BSC alone group: 50.7 vs. 108.7 days for the Panitumumab plus BSC group) - see Figure 5.1.

Comparison of responders rate based on target-lesions total size only (vs. OR metric) across the two treatment groups, adjusted for ECOG and KRAS c12/13 status provides additional evidence of response heterogeneity (p-value <.05 in both Mantel-Haenszel tests). No statistically significant difference was observed in the change of target-lesions total size, however, when comparing the same within KRAS negative status only patients, there is evidence of statistically significant lower mean increase of target-lesions total size for the experimental vs the control arm (+32.4mm vs. +45.0mm, p-value=0.036). Exploration of the individual range of response measured by target-lesions size changes in the two groups confirms that the experimental group experienced lower increase vs. that of the BSC alone group (ANOVA p-value=0.01). Calculation of the inter-individual difference in magnitude of response by the ($SD_{IR}$) metric provides evidence that heterogeneity due to treatment effect (measured by the change in target lesions total size over time) is larger than that attributed to random within-subject variability over time and has a clinically important effect size of +3.15cm. Strong evidence of responders rate difference in the two groups based on percentage of patients with at least one event of new lesion(s) during study was obtained via the generalized Cochran-Mantel-Haenszel test adjusted for the prognostic factors of ECOG and KRAS c12/13 status at baseline (p-value < .05 in both cases). Statistically significant difference is also recorded in the average number of days to new lesion(s) event between the two groups (BSC alone mean = 74.2 vs. Panitumumab plus BSC mean = 117.6 days, p-value <.001). This difference is further exaggerated within the KRAS wild-type sub-populations of the treatment groups (BSC alone mean = 44.9 vs. Panitumumab plus BSC mean = 106.4 days, p-value <.001). Figure 5.2 shows the respective distribution plots.

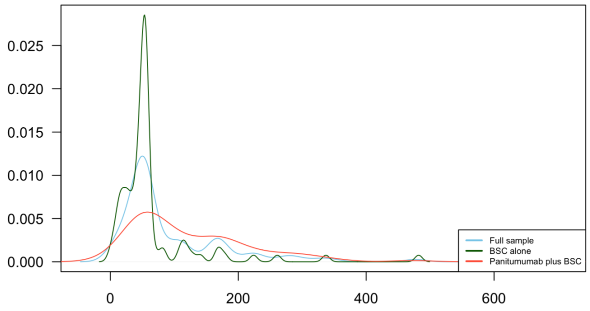


(a) Days to disease progression or death within KRAS wild-type status patients

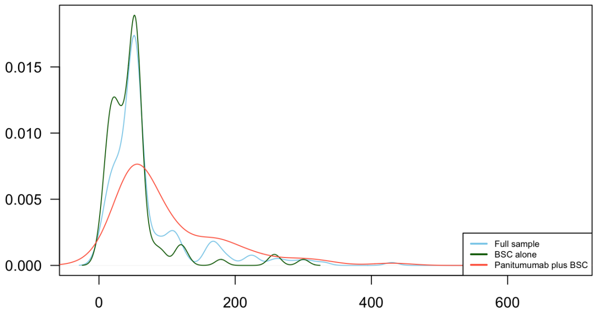


(b) Days to disease progression or death within ECOG 1 or 2 baseline status patients

Figure 5.1: Distribution plots of PFS time response metrics across treatment groups stratified by KRAS codon 12/13 negative and ECOG 1 or 2 baseline status

Table 5.1: Demographics and baseline characteristics of study population by treatment allocation

| Characteristic | Best Supportive Care (BSC) alone (N=166) Number (%) or Mean (SD) | Panitumumab Plus BSC (N=160) Number (%) or Mean (SD) | P-value |
|---|---|---|---|
| Age | 60.87 (10.34) | 61.04 (9.64) | 0.878 |
| Sex | | | 0.504 |
| *Female* | 64 (38.6) | 55 (34.4) | |
| *Male* | 102 (61.4) | 105 (65.6) | |
| Race/Ethnicity | | | 1 |
| *White* | 165 (99.9) | 160 (100.0) | |
| *Other* | 1 (0.01) | 0 (0.0) | |
| Had previous surgery | | | 1 |
| *No* | 4 ( 2.4) | 4 ( 2.5) | |
| *Yes* | 162 (97.6) | 156 (97.5) | |
| Months since diagnosis | 32.16 (19.06) | 30.93 (18.08) | 0.552 |
| Baseline Weight (kg) | 75.12 (16.01) | 73.45 (15.27) | 0.337 |
| Baseline ECOG status† | | | 0.022 |
| *Fully active* | 59 (35.5) | 80 (50.0) | |
| *Symptoms but ambulatory* | 86 (51.8) | 68 (42.5) | |
| *In bed < 50% of the time* | 21 (12.7) | 12 ( 7.5) | |
| Histological Subtype | | | 0.075 |
| *Mucinous* | 22 (13.3) | 13 ( 8.1) | |
| *Other* | 13 ( 7.8) | 23 (14.4) | |
| *No Subtype* | 131 (78.9) | 124 (77.5) | |
| Primary diagnosis | | | 0.809 |
| *Colon* | 110 (66.3) | 103 (64.4) | |
| *Rectal* | 56 (33.7) | 57 (35.6) | |
| Biomarker KRAS (c12/13) | | | 0.113 |
| *Mutant* | 67 (40.4) | 55 (34.4) | |
| *Wild-type* | 91 (54.8) | 88 (55.0) | |
| *Failure* | 8 ( 4.8) | 17 (10.6) | |
| Baseline CEA (ng/ml) | 692.81 (1823.04) | 678.17 (1284.83) | 0.933 |
| Baseline LDH (U/L) | 499.90 (542.58) | 477.60 (510.06) | 0.702 |
| Baseline ALP (U/L) | 233.11 (204.92) | 227.79 (234.14) | 0.827 |
| Baseline Albumin (g/L) | 39.36 (5.13) | 39.33 (4.02) | 0.943 |
| Baseline Creatinine (umol/L) | 71.39 (22.01) | 68.95 (20.24) | 0.299 |
| Baseline Haemoglobin (g/L) | 126.46 (17.84) | 124.30 (16.29) | 0.255 |
| Baseline Platelets ($10^9/L$) | 267.36 (112.94) | 256.59 (93.09) | 0.348 |
| Baseline WBC ($10^9/L$) | 8.39 (3.25) | 8.14 (3.03) | 0.475 |
| Baseline target-lesions size (mm) | 190.96 (87.87) | 194.32 (95.33) | 0.742 |
| Baseline non-target lesions (units) | 5.91 (2.83) | 6.14 (2.38) | 0.419 |
| Days to death event* | 260.63 (193.62) | 260.69 (187.59) | 0.998 |
| Death event* | | | 0.204 |
| *No* | 11 ( 6.6) | 18 (11.2) | |
| *Yes* | 155 (93.4) | 142 (88.8) | |
| Progression Free Survival time† | 67.91 (84.03) | 108.84 (95.34) | <0.001 |
| Disease progression during study | | | 0.258 |
| *No* | 5 ( 3.0) | 10 ( 6.2) | |
| *Yes* | 161 (97.0) | 150 (93.8) | |
| Objective Response OR† | | | <0.001 |
| *PD or UE* | 153 (92.2) | 141 (88.1) | |
| *PR or Stable* | 13 ( 7.8) | 19 ( 11.9) | |

Results are summarized as counts and percentages for categorical variables and as means and standard deviations for continuous numerical variables. The hypothesis of qualitative differences across characteristics was tested with chi-square test for categorical variables (with continuity correction) and regular ANOVA one-way test for continuous variables (with equal variance assumption). Exact test and non-normal tests were used where required. A P-value of <0.05 was considered statistically significant for all implementations.
*recorded during the study and up to the end of follow-up period.
† statistically significant difference at p-value <.05

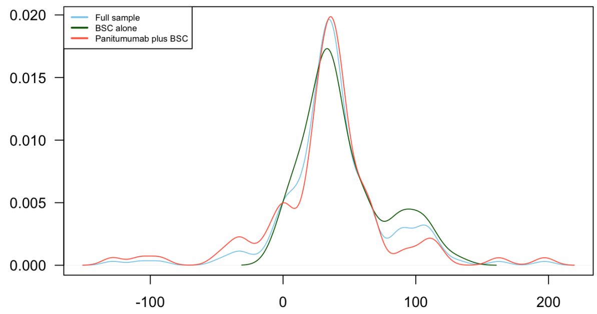


(a) Change in target-lesions total size over time (mm) within KRAS negative patients

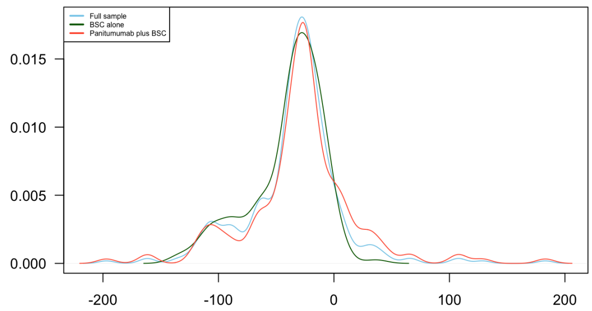


(b) Individual range of change in target-lesion total size over time (mm)

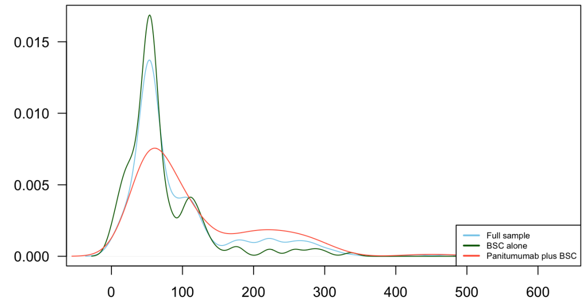


(c) Days to new lesion(s) appearance event

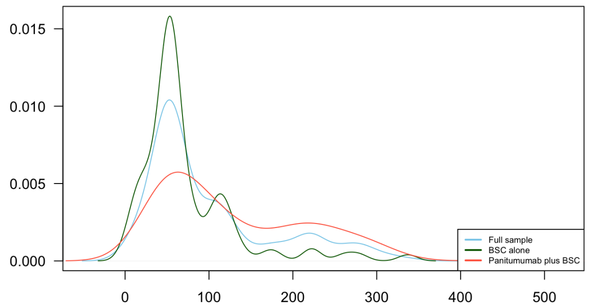


(d) Days to new lesion(s) appearance event within KRAS negative patients

Figure 5.2: Distribution plots of derived response metrics across treatment groups and their sub-populations

Further exploration of response measured by change in target-lesions total size in the BSC alone group proves the existence of outcomes heterogeneity even in absence of intervention. Figure 5.3(a) shows the two mixture densities of response metric within the BSC group with about 26% of the sub-population recording extremely negative outcomes and an average increase in tumour size over three times higher that of the rest 74% of the control population (82.6mm vs. 26.6mm). Looking into the individual range of response based on the same metric, as expected, only 31% of the BSC alone population experiences an average increase of tumour size greater than 50mm, whilst almost 70% of that population records less than half of that (Figure 5.3(b)).

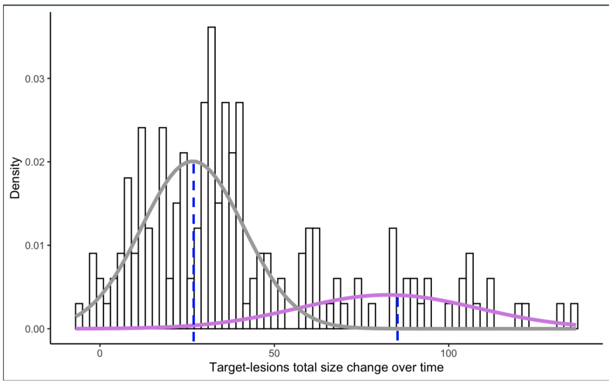


(a) Mixture distribution of change in target-lesions total size in the BSC alone treatment group

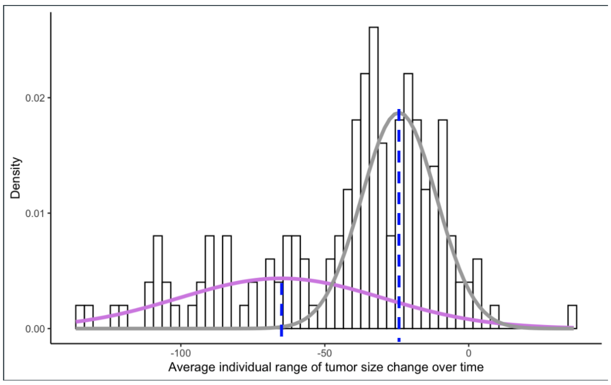


(b) BSC alone group: Mixture distribution of average individual range of tumor response by decrease in target-lesions total size

Figure 5.3: BSC alone group: Mixture distribution density plots of target-lesions total size change over time

Finally, although CEA changes over time is very low in both treatment groups, further exploration of dynamic

CEA changes during the first three months of the study and comparison of responders rate in subgroups defined by a clinically meaningful change threshold value (-50%), provide strong evidence that CEA is a sound predictor of tumour response. The rate of "responders" (OR-based) within the subgroup that achieved a decrease in CEA equal to or higher than 50% within the first 12 weeks, is more than double to that of patients who failed to reach such threshold (88% vs. 39%, p-value $<.0001$). Respective exploration of other blood markers changes over time - correlated with tumour response - namely LDH and ALP, results into similar findings.

### 3 Outcome Prediction & Heterogeneity Factors

Days to disease progression or death were statistically significant improved for the Panitumumab plus BSC group compared with the BSC alone group (p-value $<.001$, stratified log-rank test) - Figure 5.4. Median PFS time was 52 days (95% CI, 50 to 55) for BSC alone group vs. 59 days (95% CI 56 to 84) for the Panitumumab plus BSC group. The difference in median PFS time between the two groups is almost doubled

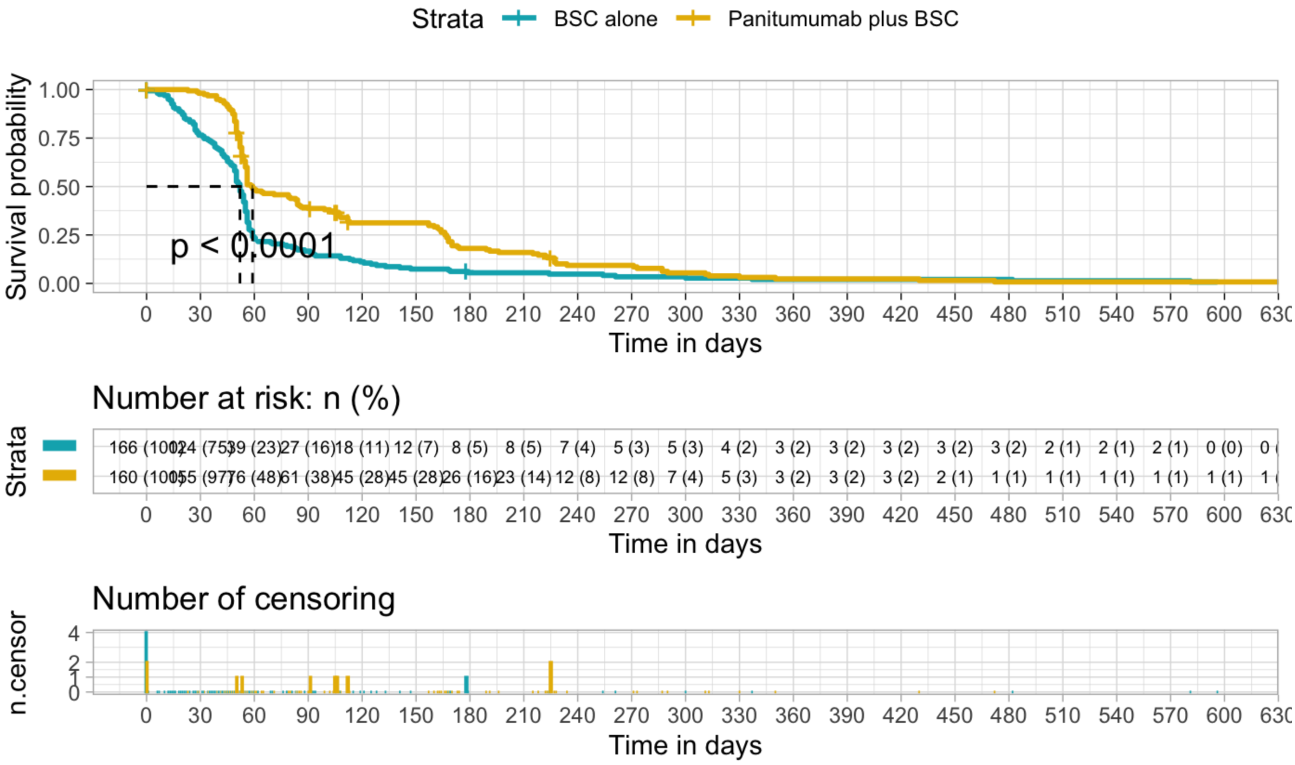


Figure 5.4: Kaplan–Meier estimates of the probability of Progression-Free Survival according to treatment arm

when further stratifying the populations for KRAS c12/13 status (52 weeks, 95% CI from 49 to 55 vs. 102 weeks, 95% CI from 79 to 164 within KRAS negative patients). The favourable effect of the interventional treatment on PFS time was consistent also after adjusting for all other variables and prognostic factors (Table 5.5). Patients who received the experimental treatment had almost half the disease progression hazard compared to that of the BSC alone treatment group (HR: 0.59, 95%CI from 0.46 to 0.76). Greater variability in target-lesions total size change favours longer PFS times (1% less hazard per mm change) suggesting that heterogeneity in response magnitude based on tumour size is an important predictor of time to disease progression regardless of the treatment allocation. Longer time since disease diagnosis at baseline also results in improved PFS times. Although the effect size is marginal per additional month, this could be associated with differential mechanisms of resistance to disease. Baseline ECOG status, other than "fully

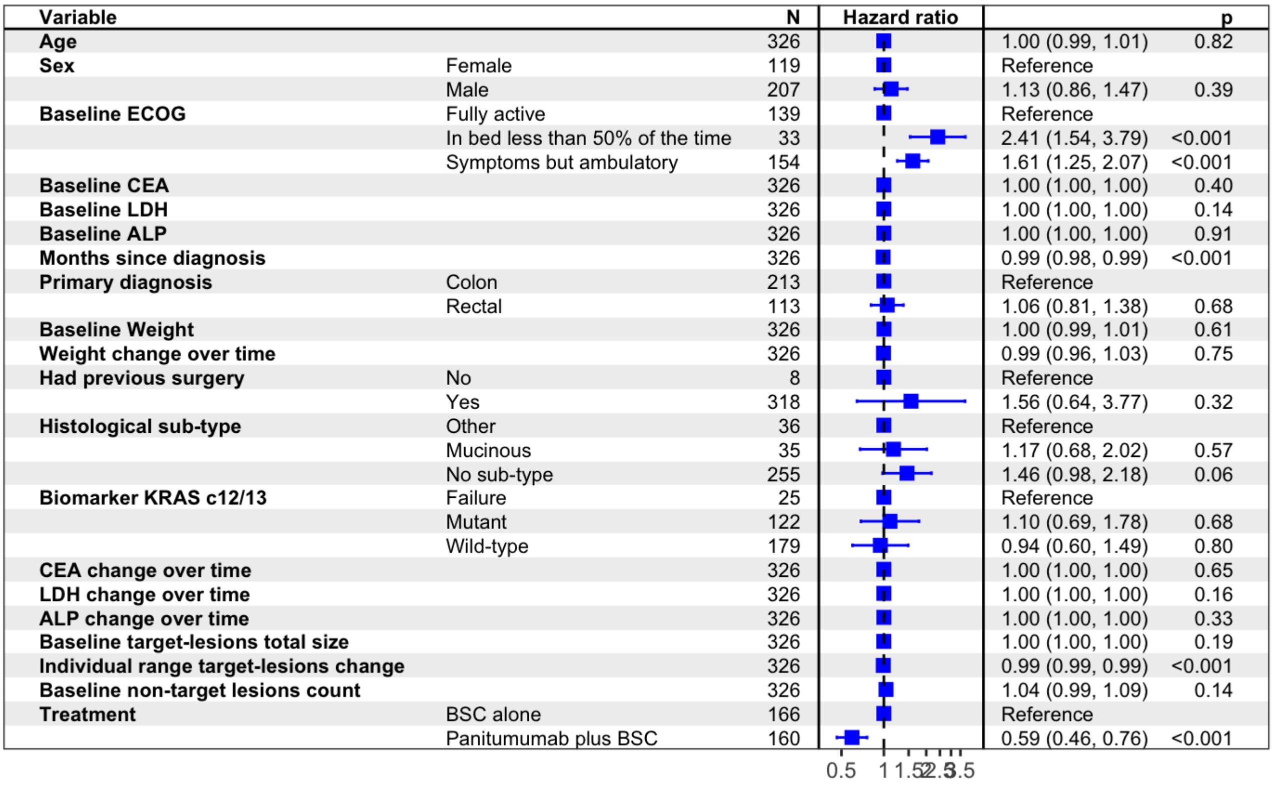


Figure 5.5: Progression-free survival (PFS) analysis: Adjusted Hazard Ratios for PFS (rectangles) and their 95% CIs (horizontal lines).

active", increases the risk for disease progression or death from 61% ("ambulatory" status) to 141% higher ("in bed < 50% of the time"). Blood marker baseline or change over time values do not impact significantly the hazard rate when adjusted for all other risk and prognostic factors.

### 4 Internal Validation

Table 5.2: Internal Validation metrics comparison across algorithms and dataset approaches

| . | of Clusters | Connectivity | Dunn's index | Avg. Silhouette Width | BIC |
|---|---|---|---|---|---|
| Extended Response data approach | | | | | |
| HAC | 3 | 11.7 | 0.1 | 0.39 | NA |
| PAM | 7 | 153.5 | 0.04 | 0.25 | NA |
| GMM | 5 | 229.9* | 0.03* | 0.04* | -2798.61 |
| GMM *with Prior* | 3 | 225.6* | 0.02* | 0.15* | -2448.54 |
| Haemetology Enriched Extended Response data approach | | | | | |
| HAC | 2 | 2.93 | 0.41 | 0.52 | NA |
| PAM | 2 | 39.53 | 0.07 | 0.22 | NA |
| GMM | 4 | 222.35* | 0.05* | 0.03* | -7271.498 |
| GMM *with Prior* | 3 | 173.81* | 0.05* | 0.08* | -5930.461 |

**Connectivity, Dunn's index and Average Silhouette Width metrics are less informative for non-linear based clustering methods.*

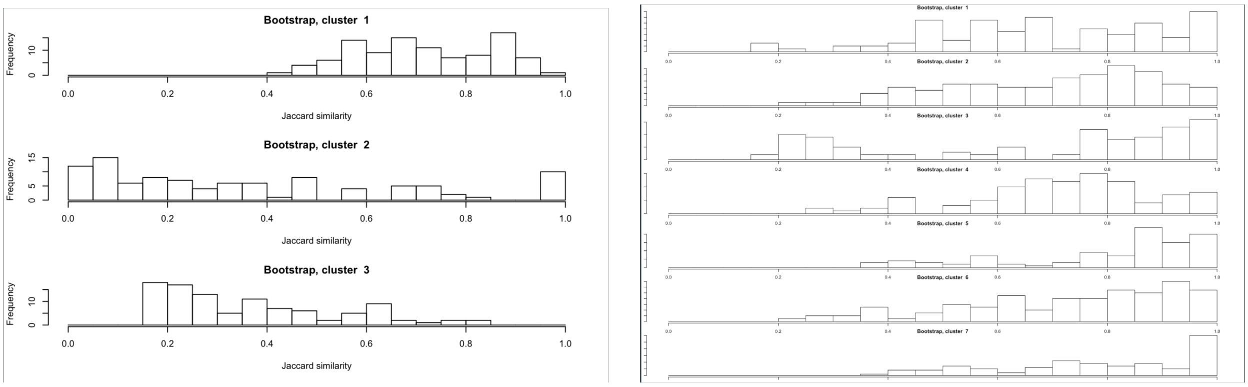


(a) Approach 2 - HAC clusterwise stability assessment

(b) Approach 2 - PAM clusterwise stability assessment

Figure 5.6: Clusterwise stability assessment by resampling

### 5 Supplementary Data - Sensitivity Analysis

Notes applicable to following tables: Results are summarized as counts and percentages for categorical variables and as means and standard deviations for continuous numerical variables. The hypothesis of qualitative differences across characteristics was tested with chi-square test for categorical variables (with continuity correction) and regular ANOVA one-way test for continuous variables (with equal variance assumption). Exact test and non-normal tests were used where required. A P-value of $<0.05$ was considered statistically significant for all implementations.
*recorded during the study and up to the end of follow-up period.
† statistically significant difference at p-value $<.05$

Table 5.3: Baseline, outcome & response characteristics of interventional arm patient subgroups based on OR

| Characteristic | Non-responders: PD or UE (N=138) | Responders: PR or Stable (N=22) | P-value |
|---|---|---|---|
| Age | 61.07 (9.55) | 60.86 (10.42) | 0.933 |
| Sex | | | 0.349 |
| *Female* | 45 ( 32.6) | 10 ( 45.5) | |
| *Male* | 93 ( 67.4) | 12 ( 54.5) | |
| Had previous surgery | | | 0.45 |
| *No* | 3 ( 2.2) | 1 ( 4.5) | |
| *Yes* | 135 ( 97.8) | 21 ( 95.5) | |
| Months since diagnosis | 30.38 (18.27) | 34.37 (16.80) | 0.315 |
| Baseline Weight (kg) | 74.03 (15.36) | 69.79 (14.48) | 0.215 |
| Baseline ECOG status | | | 0.435 |
| *Fully active* | 69 ( 50.0) | 11 ( 50.0) | |
| *Symptoms but ambulatory* | 60 ( 43.5) | 8 ( 36.4) | |
| *In bed < 50% of the time* | 9 ( 6.5) | 3 ( 13.6) | |
| Histological Subtype | | | 0.587 |
| *Mucinous* | 10 ( 7.2) | 3 ( 13.6) | |
| *Other* | 20 ( 14.5) | 3 ( 13.6) | |
| *No Subtype* | 108 ( 78.3) | 16 ( 72.7) | |
| Primary diagnosis | | | 0.521 |
| *Colon* | 87 ( 63.0) | 16 ( 72.7) | |
| *Rectal* | 51 ( 37.0) | 6 ( 27.3) | |
| Biomarker KRAS exon 2(c12/13) | | | 0.419 |
| *Mutant* | 50 ( 36.2) | 5 ( 22.7) | |
| *Wild-type* | 74 ( 53.6) | 14 ( 63.6) | |
| *Failure* | 14 ( 10.1) | 3 ( 13.6) | |
| Baseline CEA (ng/ml) | 688.15 (1236.80) | 615.52 (1585.80) | 0.839 |
| Baseline LDH (U/L) | 478.84 (502.56) | 469.82 (567.45) | 0.944 |
| Baseline ALP (U/L) | 229.72 (245.54) | 215.68 (146.89) | 0.711 |
| Baseline Albumin (g/L) | 39.44 (3.89) | 38.59 (4.81) | 0.437 |
| Baseline Creatinine (umol/L)† | 70.32 (20.95) | 60.36 (12.31) | 0.003 |
| Baseline Hemoglobin (g/L) | 124.63 (16.51) | 122.24 (14.99) | 0.499 |
| Baseline Platelets ($10^9/L$) | 263.01 (94.22) | 216.32 (75.71) | 0.014 |
| Baseline WBC ($10^9/L$) | 8.24 (3.18) | 7.49 (1.73) | 0.106 |
| Baseline target-lesions size (mm) | 193.68 (96.03) | 198.31 (92.91) | 0.831 |
| Baseline non-target lesions (units) | 6.25 (2.45) | 5.45 (1.82) | 0.078 |
| Weight change (kg)* | -0.99 (3.78) | -0.85 (2.96) | 0.835 |
| CEA change (ug/L)* | 282.76 (1467.63) | 105.86 (1060.81) | 0.498 |
| LDH change (U/L)*† | 186.46 (624.26) | -45.82 (198.80) | 0.001 |
| ALP change (U/L)* | 80.54 (198.43) | 22.05 (173.10) | 0.16 |
| Objective Response OR† | | | < 0.001 |
| *PD or UE* | 138 (100.0) | 3 (13.6) | |
| *PR or Stable* | 0 (0.0) | 19 (86.4) | |
| Objective Response - Target lesions† | | | < 0.001 |
| *PD or UE* | 93 (67.4) | 3 (13.6) | |
| *PR or Stable* | 45 (32.6) | 19 (86.4) | |
| Target-lesions size change*† | 43.89 (37.15) | -6.90 (49.31) | <0.001 |
| Target-lesions max size (mm)* | 246.44 (116.32) | 220.03 (102.32) | 0.279 |
| Target-lesions max increase (mm)*† | 51.82 (33.27) | 19.71 (23.23) | <0.001 |
| Target-lesions size change rate*† | 0.23 (0.22) | -0.05 (0.22) | <0.001 |
| Target-lesions min size (mm)* | 182.97 (96.47) | 166.94 (87.63) | 0.438 |
| Target-lesions min size day† | 26.83 (48.98) | 57.77 (53.99) | 0.018 |
| Ind. Range of target-lesions response (mm)*† | -35.34 (43.13) | 14.24 (57.27) | 0.001 |
| Days to PD based on target-lesions size | 153.28 (104.86) | 155.27 (94.75) | 0.929 |
| New lesions event* | | | 0.528 |
| *No* | 4 ( 2.9) | 1 ( 4.5) | |
| *Yes* | 134 ( 97.1) | 21 ( 95.5) | |
| New lesions size last visit (mm)† | 28.19 (20.33) | 18.76 (9.11) | 0.001 |
| New lesions event day*† | 102.88 (69.82) | 209.64 (119.91) | <0.001 |
| Days to Death event† | 248.64 (176.50) | 336.23 (237.21) | 0.109 |
| Death event* | | | 0.078 |
| *No* | 13 ( 9.4) | 5 ( 22.7) | |
| *Yes* | 125 ( 90.6) | 17 ( 77.3) | |
| Progression Free Survival time*† | 96.96 (82.78) | 183.36 (131.81) | 0.007 |
| Disease Progression event*† | | | <0.001 |
| *No* | 0 ( 0.0) | 10 ( 45.5) | |
| *Yes* | 138 (100.0) | 12 ( 54.5) | |

Table 5.4: Baseline, outcome & response characteristics of interventional arm patient subgroups: PAM membership

| Characteristic | Cluster 1 (N=63) | Cluster 2 (N=39) | Cluster 3 (N=58) | P-value |
|---|---|---|---|---|
| Age | 60.65 (8.09) | 62.56 (10.90) | 60.43 (10.33) | 0.585 |
| Sex | | | | 0.852 |
| *Female* | 20 ( 31.7) | 14 ( 35.9) | 21 ( 36.2) | |
| *Male* | 43 ( 68.3) | 25 ( 64.1) | 37 ( 63.8) | |
| Had previous surgery | | | | 0.832 |
| *No* | 1 ( 1.6) | 1 ( 2.6) | 2 ( 3.4) | |
| *Yes* | 62 ( 98.4) | 38 ( 97.4) | 56 ( 96.6) | |
| Months since diagnosis | 32.62 (15.79) | 33.01 (14.58) | 27.70 (21.96) | 0.301 |
| Baseline Weight (kg) | 72.56 (15.60) | 75.83 (14.49) | 72.82 (15.51) | 0.514 |
| Baseline ECOG status | | | | 0.18 |
| *Fully active* | 29 ( 46.0) | 24 ( 61.5) | 27 ( 46.6) | |
| *Symptoms but ambulatory* | 26 ( 41.3) | 13 ( 33.3) | 29 ( 50.0) | |
| *In bed less than 50% of the time* | 8 ( 12.7) | 2 ( 5.1) | 2 ( 3.4) | |
| Histological Subtype | | | | 0.356 |
| *Mucinous* | 3 ( 4.8) | 2 ( 5.1) | 8 ( 13.8) | |
| *Other* | 10 ( 15.9) | 7 ( 17.9) | 6 ( 10.3) | |
| *No Subtype* | 50 ( 79.4) | 30 ( 76.9) | 44 ( 75.9) | |
| Primary diagnosis | | | | 0.71 |
| *Colon* | 43 ( 68.3) | 24 ( 61.5) | 36 ( 62.1) | |
| *Rectal* | 20 ( 31.7) | 15 ( 38.5) | 22 ( 37.9) | |
| Biomarker KRAS exon 2(c12/13)† | | | | 0.001 |
| *Mutant* | 22 ( 34.9) | 4 ( 10.3) | 29 ( 50.0) | |
| *Wild-type* | 35 ( 55.6) | 29 ( 74.4) | 24 ( 41.4) | |
| *Failure* | 6 ( 9.5) | 6 ( 15.4) | 5 ( 8.6) | |
| Baseline CEA (ng/ml) | 827.78 (1728.59) | 607.97 (947.53) | 562.86 (845.30) | 0.559 |
| Baseline LDH (U/L) | 569.02 (597.74) | 396.08 (291.96) | 433.12 (514.25) | 0.155 |
| Baseline ALP (U/L) | 234.13 (231.31) | 231.10 (207.08) | 218.67 (256.90) | 0.938 |
| Baseline Albumin (g/L) | 38.57 (4.59) | 39.56 (3.20) | 39.98 (3.77) | 0.18 |
| Baseline Creatinine (umol/L) | 67.76 (20.85) | 67.21 (14.07) | 71.41 (22.99) | 0.517 |
| Baseline Hemoglobin (g/L) | 122.69 (15.38) | 127.05 (14.36) | 124.21 (18.37) | 0.355 |
| Baseline Platelets ($10^9/L$) | 249.52 (92.91) | 243.21 (89.96) | 273.26 (94.48) | 0.227 |
| Baseline WBC ($10^9/L$) | 8.43 (3.48) | 7.35 (2.02) | 8.35 (3.03) | 0.064 |
| Baseline target-lesions size (mm) | 195.22 (109.80) | 195.77 (65.22) | 192.36 (96.94) | 0.978 |
| Baseline non-target lesions (units)† | 6.92 (2.59) | 5.79 (1.54) | 5.53 (2.41) | 0.006 |
| Weight change* (kg) | -1.24 (3.85) | 0.21 (4.00) | -1.47 (3.08) | 0.085 |
| CEA change (ng/ml)* | 175.93 (1567.92) | 53.66 (1457.94) | 485.75 (1192.54) | 0.241 |
| LDH Change (U/L)* | 131.83 (693.43) | 91.85 (437.22) | 221.33 (557.58) | 0.437 |
| ALP Change (U/L)*† | 57.22 (176.20) | 21.90 (136.72) | 123.11 (236.58) | 0.033 |
| Objective Response OR† | | | | 0.001 |
| *PD or UE* | 50 (79.4) | 33 (84.6) | 58 (100.0) | |
| *PR or Stable* | 13 (20.6) | 6 (15.4) | 0 (0.0) | |
| Objective Response - Target lesions† | | | | <0.001 |
| *PD or UE* | 9 (14.3) | 29 (74.4) | 58 (100.0) | |
| *PR or Stable* | 54 (85.7) | 10 (25.6) | 0 (0.0) | |
| Target-lesions size change (mm)*† | 22.49 (34.06) | 18.24 (36.41) | 65.13 (40.59) | <0.001 |
| Target-lesions max size (mm)* | 229.51 (125.94) | 238.20 (73.09) | 260.36 (123.57) | 0.373 |
| Target-lesions size max increase*† | 33.93 (26.07) | 38.94 (21.09) | 67.74 (38.69) | <0.001 |
| Target-lesions size change rate*† | 0.09 (0.18) | 0.10 (0.19) | 0.37 (0.22) | <0.001 |
| Target-lesions min size (mm)* | 183.50 (110.90) | 162.28 (58.93) | 190.21 (96.22) | 0.169 |
| Target-lesions min size day† | 29.90 (38.85) | 86.00 (48.17) | -4.55 (24.75) | <0.001 |
| Ind. Range of target-lesions response (mm)*† | -17.41 (32.66) | 4.16 (47.97) | -62.57 (41.92) | <0.001 |
| Days to PD based on target-lesions size† | 117.37 (56.70) | 298.18 (95.47) | 95.62 (31.43) | <0.001 |
| New lesion(s) event* | | | | 0.253 |
| *No* | 1 ( 1.6) | 3 ( 7.7) | 1 ( 1.7) | |
| *Yes* | 62 ( 98.4) | 36 ( 92.3) | 57 ( 98.3) | |
| New lesion(s) size last visit | 28.17 (19.77) | 25.64 (19.33) | 26.34 (19.39) | 0.791 |
| New lesion(s) event day† | 97.98 (75.85) | 225.49 (66.56) | 66.26 (22.86) | <0.001 |
| Days to Death event† | 222.49 (168.25) | 458.77 (187.57) | 168.98 (83.09) | <0.001 |
| Death event* | | | | 0.505 |
| *No* | 5 ( 7.9) | 6 ( 15.4) | 7 ( 12.1) | |
| *Yes* | 58 ( 92.1) | 33 ( 84.6) | 51 ( 87.9) | |
| Progression Free Survival time*† | 83.92 (74.89) | 222.87 (100.71) | 59.22 (20.17) | <0.001 |
| Disease Progression event*† | | | | 0.009 |
| [illegible] | [illegible] | [illegible] | [illegible] | |
